# Removal of the race coefficient and adjustment to individual BSA provide the most accurate estimation for GFR in Black adolescents

**DOI:** 10.1101/2021.07.30.21261371

**Authors:** D Bielopolski, O Bentur, N Singh, R Vaughan, DM Charytan, RG Kost, JN Tobin

## Abstract

**Introduction:** Obesity is more prevalent among minorities, increasing the risk for cardio-renal morbidity. We explored interactions between race, body mass index (BMI), and the risk of hyperfiltration associated with Obesity Related Glomerulopathy (ORG).

**Methods:** We created a cohort of women and girls ages 12-21 from the New York Metropolitan area using electronic health records. Glomerular filtration rate (GFR) was estimated in three ways: I) using the standard age recommended formulae, II) eGFRr – without a race-specific coefficient, and III) Absolute eGFRr – combing removal of the race coefficient and adjusting to individual body surface area. Multivariate logistic regression was used to analyze the relative contribution of risk factors for ORG associated hyperfiltration, defined by a threshold of ≥135ml/min/1.73m^2^. Bland Altman analysis and Pearson’s coefficient assessed the correlation of each formula with creatinine clearance (CrCl).

**Results:** 7315 Black and 15,102 non-Black women and girls had simultaneous evaluation of kidney function and body measures. CrCl was available in 207 non-Black and 107 Black individuals as an internal validation. Simultaneous removal of the race coefficient and adjustment to individual BSA estimated GFR most accurately compared to CrCl, across BMI groups and between races. Hyperfiltration was more frequent in obese Black compared to non-Black individuals when using standard eGFR (20% vs. 6.5% respectively) but had a lower frequency after eliminating the race-specific coefficient (4.5% vs. 6.5%). Black race was independently associated with a higher risk of hyperfiltration with standard eGFR calculations (OR=3.43, 95% CI 2.95-3.99) and with lower risk when estimated by eGFRr (OR=0.56, 95% CI 0.45-0.70). Simultaneous removal of the race coefficient from GFR calculation and adjustment to individual BSA attenuated the difference in risk between races (OR=0.8, 95% CI 0.68-0.94). The combined correction agreed well with creatinine clearance (Pearson’s correlation coefficient r=0.64, 0.52 and 0.52 for absolute eGFRr, eGFR and eGFRr respectively.

**Conclusions:** Removal of the race coefficient from GFR estimating equations obscures obesity associated hyperfiltration among Black adolescents. This correction should be accompanied by adjustment to individual BSA to improve estimation of GFR to avoid misclassification of obesity related hyperfiltration.

## Introduction

Black individuals are at higher risk of kidney diseases(1,2) accounting for one third of all kidney failures in the United Sates despite accounting for only 14% of the population(1). Decline in GFR among Black individuals occurs at an earlier age and at a faster annualized rate when compared with White Individuals, even by cystatin C–based GFR assessment(3).

This may partly relate to the disproportionately high rates of obesity in ethnic minority, low-income, and other socially marginalized US population groups. The prevalence of obesity is significantly higher in non-Hispanic Black (55%) and Hispanic (51%) than non-Hispanic White women (38%)(4), and obesity is associated with increased glomerular filtration rate (GFR), also known as hyperfiltration(5), and glomerular structural changes, such as mesangial matrix expansion, and abnormal renal hemodynamics that are reversible in early stages(6,7).

Thus, hyperfiltration in obese patients might be viewed as an early marker of kidney dysfunction, similar to glomerular hyperfiltration in patients with diabetes or hypertension that precedes the development of microalbuminuria(8,9) eventually leading to irreversible focal glomerulosclerosis(10) and the need for renal replacement therapy(11).

Estimation of eGFR is required to identify hyperfiltration as well as the presence of CKD in clinical practice, but typically relies on serum-creatinine based equations incorporating use of a race coefficient to separately estimate eGFR in Black and non-Black individuals.(12). Use of the race coefficient has been questioned by members of the nephrology community, since it has been shown to postpone timely patient referral to specialist evaluation, dialysis and transplantation (13–16). Yet, eliminating the coefficient could lead to unintended consequences such as overdiagnosis of CKD, inadequate use or dosing of drugs excreted by glomerular filtration (including metformin), and limited access to tests (including imaging procedures) and treatments that require a higher level of GFR(13). To the best of our knowledge, however, the impact of race-specific eGFR coefficient on identification of hyperfiltration, the earliest manifestation of obesity-related glomerulopathy (ORG) has not been evaluated.

Following the initiative to replace race-based equations by a substitute that is accurate, representative, and unbiased(17), we sought to analyze whether elimination of race-based equations could affect identification of ORG among minorities and to determine if race modifies the association of BMI with the risk of hyperfiltration.

## Methods

### Cohort construction

De-identified electronic health record data were extracted for female adolescents aged 12-21 years who received health care services from 1/1/2011 to 12/31/2015 in New York City (NYC) in 12 academic health centers and federally qualified health centers(FQHCs) that are part of the Patient Centered Outcomes Research (PCOR)-funded NYC Clinical Data Research Network (NYC-CDRN, now INSIGHT)(18).

### Data-Cleaning steps for biologically plausible limits

The first encounter for each individual in which height, weight, blood pressure and serum creatinine were available was included. Extreme outlier values that might indicate data entry errors rather than true outliers were excluded by setting physiological limits for systolic blood pressure (60-220 mmHg), diastolic blood pressure (30-150 mmHg), BMI (12-80 kg/m^2^), height (127-200cm), weight (24-240 kg) and serum creatinine (0.3-3 mg/dL).

BMI was calculated by dividing weight in kilograms by the square of height in meters. BMI values were classified according to the Centers for Disease Control and Prevention (19) data for BMI-for-age z-score into the following categories: Underweight: BMI ≤5^th^ percentile, Overweight: 95^th^≥BMI≥85^th^ percentile, Obese: BMI ≥95^th^ percentile, all for children and teens of the same age and sex.(19) BSA was calculated using the metric system according to the Du-Bois formula (*BSA* = 0.007184 * Height^0.725^ * Weight^0.425^)(20).

The cohort included adolescents from multiple races, for whom renal function were documented. Since the race coefficient relates to Black individuals in comparison to non-Black individuals(12) we divided the cohort accordingly. As part of the efforts to ascertain best clinical practice, all patients were asked to self-define their race and ethnicity.

### eGFR calculation

We calculated eGFR using 3 different formulae: 1) KDIGO recommended for age formulae(21), using CKD-EPI for 18≤21 year old, and Schwartz formulae for 12-18 year old, 2) eGFRr was calculated without a race specific coefficient which applied only for Blacks 18–21 years old, as the Schwartz formula does not contain a race coefficient. 3) Absolute eGFRr - calculated without a race specific coefficient and adjusted to individual BSA.Hyperfiltration was defined according to published criteria as BSA-standardized eGFR >135 mL/min/1.73 m^2^ (22).

### Statistical Analysis

Statistical analyses were performed using SAS (r 9.4). Nominal variables were expressed as numbers (%). Comparison of proportions between groups was performed using the chi-squared test. Continuous variables were expressed as mean ± SD and were compared using ANOVA including Dunnett’s approach for multiple comparisons, where a p<0.05 (two-tailed) was considered statistically significant.

Regression model - Univariable analyses were used to describe associations between the different variables and hyperfiltration. Multivariate logistic regression models for hyperfiltration used the predictor variables as fixed effects. These included: age, BMI, systolic and diastolic blood pressure, glycosylated Hemoglobin (HbA1c), and race. All effect estimates are presented as odds ratio (OR), along with the corresponding 95% confidence intervals (CI).

Bland Altman analysis - Bland-Altman’s analysis was used to assess the relative agreement between creatinine clearance, eGFR and absolute eGFRr. Pearson’s coefficient was used to assess correlation between continuous variables.

Ethics declaration - Data extraction and transmission were reviewed and approved, and a waiver of informed consent for analyzing de-identified data was granted by the Institutional Review Boards at Clinical Directors Network (CDN), BRANY (Biomedical Research Alliance of New York), and the Rockefeller University.

## Results

### Cohort construction

The cohort included 22,417 unique female patients with serum creatinine values recorded, of whom 7,315 individuals (32.6%) were Black (Table 1). A higher proportion of Black subjects (26%) were obese compared to 20% of non-Black individuals (Table 1). Blood pressure increased across BMI in both Black and White individuals, as previously described(23).

**Table 1.** Demographic and vital characteristics of Black individuals vs. non-Black individuals (22,417 patients).

|  | Non-Black<br>(15,102) |  |  |  | Blacks<br>(7,315) |  |  |  |
| --- | --- | --- | --- | --- | --- | --- | --- | --- |
|  | Underweight<br>(786) | Normal<br>(8,340) | Overweight<br>(2,886) | Obese<br>(3,090) | Underweight<br>(299) | Normal<br>(3,631) | Overweight<br>(1,467) | Obese<br>(1,918) |
| Age<br>(years) | 16.9±2.8 | 17.1±2.5 | 17.3±2.5 | 17.1±2.5 | 17.12±2.9 | 17.14±2.5 | 17.35±2.6 | 17.14±2. |
| BSA (m<sup>2</sup>) | 1.3±0.2 | 1.5±0.2 | 1.7±0.1 | 1.9±0.2 | 1.4±0.2 | 1.6±0.2 | 1.8±0.2 | 2±0.2 |
| Systolic BP<br>(mmHG) | 102±11 | 105±11 | 110±11 | 114±12 | 103±12 | 106±11 | 111±12 | 116±13 |
| Diastolic<br>BP(mmHG) | 64±8 | 64±8 | 66±8 | 68±9 | 63±9 | 65±8 | 67±9 | 70±10 |
| HbA1c (%) | 5.6 | 5.6 | 5.6 | 5.6 | 5.8 | 5.6 | 5.7 | 5.6 |

### Validation cohort

CrCl data from 24-hour urine collections were available for 309 individuals of whom 102 (33%) were black. CrCl increased across BMI groups for both races, of Blacks and non-Black individuals, and were statistically similar for the overweight and obese (Mean CrCL for overweight Black 132ml/min vs. overweight non-Black 130ml/min, obese Black 138ml/min vs. obese non-Black 142ml/min individuals). In contrast, eGFR values were higher for Black individuals compared to non-Blacks in every BMI group, but this pattern was reversed for the overweight and obese when using eGFRr (overweight and obese Black; 111±31, 113±31, vs. overweight and obese non-Black; 126±17, 119±21 respectively).

GFR estimation was closest to creatinine clearance when using absolute eGFRr, combing both race coefficient removal and adjustment to individual BSA, for Black individuals which was consistent across all BMI groups (Table 2 and Figure 1). Comparing absolute eGFRr for Black individuals with absolute eGFR for non-Black eliminated the differences between racial groups, across all BMI subgroups (table 2).

**Table 2.** Renal characteristics of Black individuals vs. non-Black individuals in cohort of 309 individuals with available clearance data. eGFR – calculated according to KDIGO recommended for age formulae, eGFRr – eGFR after removal of race correction factor, applicable only for the black patients ages 18-21. For non-Black individuals values represent estimation without manipulation of race coefficient i.e instead of eGFRr>eGFR, Absolute eGFRr>Absolute eGFR. P-values for creatinine clearance subgroups represent comparison across races for these subgroups.

|  | Non-Black<br>(207) |  |  |  | Blacks<br>(102) |  |  |  |
| --- | --- | --- | --- | --- | --- | --- | --- | --- |
|  | Underweight<br>(6) | Normal<br>(89) | Overweight<br>(39) | Obese<br>(73) | Underweight<br>(4) | Normal<br>(35) | Overweight<br>(23) | Obese<br>(40) |
| Serum Creatinine (mg/dl) | 0.71 | 0.68 | 0.60 | 0.63 | 0.65 | 0.68 | 0.80 | 0.70 |
| Creatinine clearance (ml/min) | 48±13 | 104±53 | 129±53 | 142±52 | 71±28 | 116±56<br>p-value=0.67 | 132±62<br>p-value=0.42 | 138±66<br>p-value=0.07 |
| eGFR (ml/min/1.73m<sup>2</sup>) | 100±16 | 117±26 | 126±17 | 119±21 | 108±19 | 134±33 | 125±37 | 125±36 |
| eGFRr – (ml/min/1.73m<sup>2</sup>) without race coefficient | 100±16 | 117±26 | 126±17 | 119±21 | 104±17 | 121±31 | 111±31 | 113±31 |
| Absolute eGFRr (ml/min) without race coefficient) | 73±22 | 104±27 | 126±17 | 134±28 | 74±16 | 111±29 | 118±37 | 133±40 |

**Figure 1.**
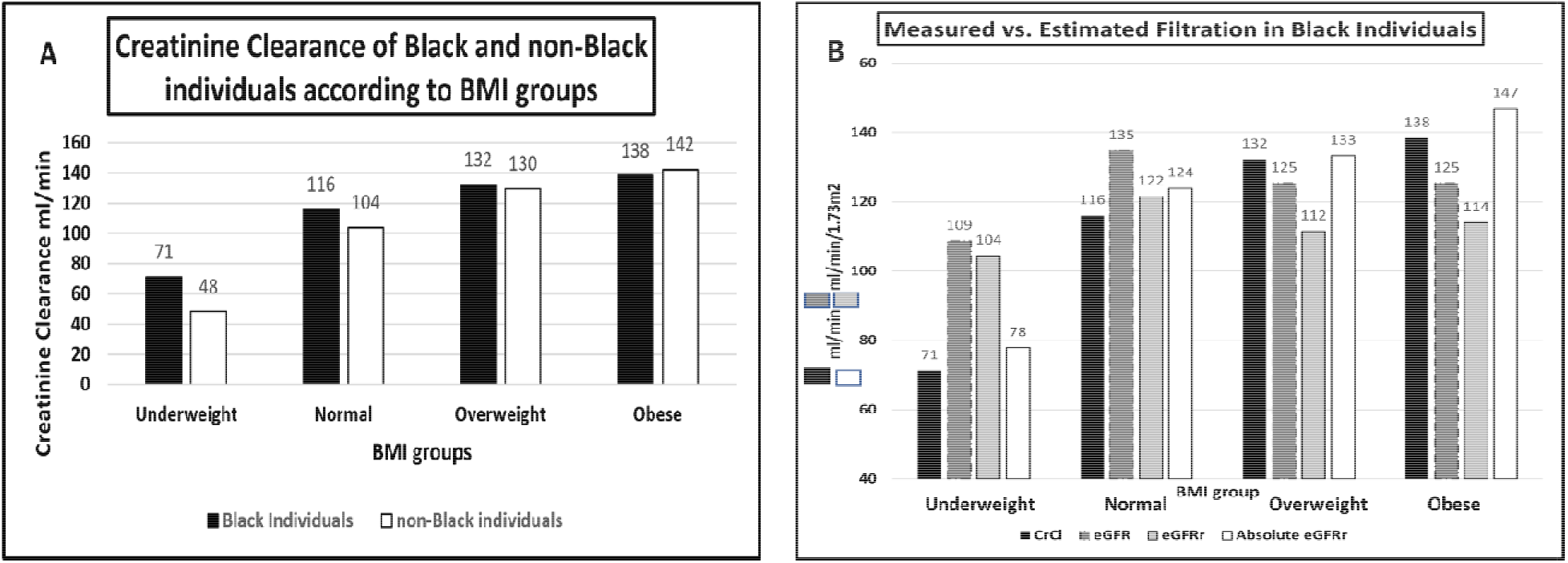
Measured vs. estimated GFR according to race in 309 individuals with available creatinine clearance data. Panel A shows measured creatinine clearance according to race and BMI groups. Panel B shows measure GFR vs. different estimation formulae in Black individuals.

### Renal characteristics for the full cohort

Using the standard calculation of eGFR, mean eGFR values were significantly higher across all BMI groups for Black individuals compared to non-Black individuals (Table 3). However, after eliminating the race-specific coefficient, eGFRr values were significantly lower between groups for the obese patients (119±21 ml/min/1.73m^2^ for obese non-Black individuals vs. 113±31 ml/min/1.73m^2^ for obese black individuals), although the difference was marginal. Estimated GFR remained higher in Black participants in the normal and underweight subgroups (Table 2) after elimination of the race-specific coefficient.

**Table 3.** Estimated GFR for 22,417 individuals according to the different formulae. Numbers in brackets are number of individuals with available data. eGFR – recommended formulae for age. eGFRr – recommended formulae after removal of the race coefficient. Applied only for Black individuals ages 18-21. Absolute eGFRr – recommended formulae after removal of the race coefficient and adjusted to individual BSA. For non-Black individuals values represent estimation without manipulation of race coefficient i.e instead of eGFRr>eGFR, Absolute eGFRr>Absolute eGFR

|  | Non-Black<br>(15,102) |  |  |  | Blacks<br>(7,315) |  |  |  |
| --- | --- | --- | --- | --- | --- | --- | --- | --- |
|  | Underweight<br>(786) | Normal<br>(8,340) | Overweight<br>(2,886) | Obese<br>(3,090) | Underweight<br>(299) | Normal<br>(3,631) | Overweight<br>(1,467) | Obese<br>(1,918) |
| Serum Creatinine (mg/dl) | 0.73 | 0.74 | 0.73 | 0.73 | 0.72 | 0.75 | 0.76 | 0.76 |
| eGFR ml/min/1.73m<sup>2</sup> | 103±29 | 101±25 | 104±25 | 104±25 | 114±32 | 109±29 | 110±29 | 107±27 |
| eGFRr – ml/min/1.73m<sup>2</sup> without race coefficient | 103±29 | 101±25 | 104±25 | 104±25 | 106±27 | 101±24 | 101±23 | 100±21 |
| Absolute eGFRr ml/min (without race coefficient) | 79±24 | 89±24 | 102±26 | 116±31 | 83±24 | 91±23 | 103±25 | 115±29 |

Adjustment to individual BSA – Absolute eGFR for the non-Black individuals was similar to absolute eGFRr for Black individuals across all BMI groups, reflecting the trend observed in the validation cohort (102±26 vs. 103±25 in the overweight, 116±31 vs. 115±29 in the obese, non-Black and Black respectively, Table 3).

The prevalence of hyperfiltration was higher among Black patients compared to non-Black patients across all BMI groups with the standard estimates of eGFR (Figure 2). However, when estimated with eGFRr, hyperfiltration was less frequent amongst Black compared with non-Black participants who were overweight or obese, while remaining more frequent for the underweight group. Combining both corrections, race coefficient removal and adjustment for individual BSA, for Black individuals, compared to individual BSA adjustment for non-Blacks, created similar hyperfiltration rates across BMI groups and between racial groups (figure 2 panel C).

**Figure 2.**
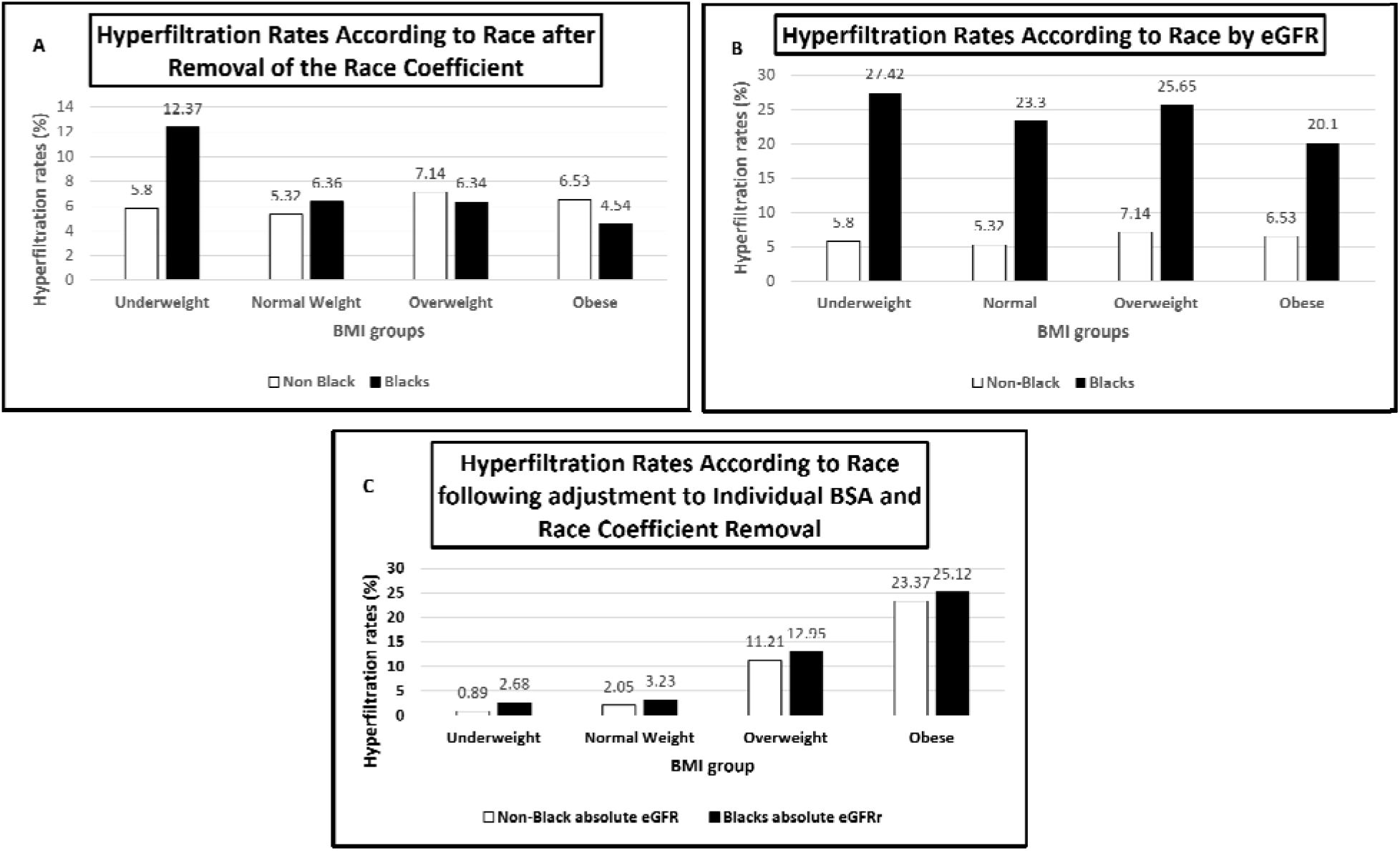
Hyperfiltration rates according to race and the different formulae in Black individuals vs. non-Black individuals in cohort of 22,417. Panel A – hyperfiltration calculated by eGFR according to BMI groups in Black and non-Black individuls. Panel B - hyperfiltration rates according to eGFRr in blacks and non-Black individuals. Panel C - hyperfiltration rates in Blacks after removal of the race coefficient and adjusting for individual BSA, and in non-Black after adjustent for individual BSA.

### Regression model

In both crude and adjusted models (Table 4), hyperfiltration according to all three formulae was significantly associated with age, and glycated hemoglobin (HbA1c). Associations of all three formulae with blood pressure parameters (either systolic or diastolic) were marginal for the crude and adjusted models.

**Table 4.** Regression model for hyperfiltration of Black individuals vs. non-Black individuals. Numbers are presented as odds ratio and 95% CI. eGFR – recommended formulae for age.eGFRr - eGFR after removal of the race coefficient. Absolute eGFRr - recommended formulae after removal of the race coefficient and adjusted to individual BSA.

| variable | eGFR |  | eGFRr |  | Absolute eGFRr |  |
| --- | --- | --- | --- | --- | --- | --- |
|  | observed | adjusted | observed | adjusted | observed | adjusted |
| Age | 1.31 (1.28-1.33) | 1.35 (1.30-1.40) | 1.09 (1.06-1.11) | 1.09 (1.05-1.14) | 1.30 (1.27-1.33) | 1.24 (1.20-1.28) |
| Systolic BP | 1.01 (1.00-1.01) | 1.00 (0.99-1.01) | 1.00 (1.00-1.01) | 1.00 (1.00-1.01) | 1.04 (1.04-1.04) | 1.02 (1.11-1.02) |
| Diastolic BP | 1.00 (1.00-1.01) | 0.99 (0.99-0.99) | 0.99 (0.99-1.00) | 0.98 (0.97-1.00) | 1.03 (1.03-1.04) | 0.99 (0.98-1.00) |
| BMI | 1.02 (1.01-1.02) | 0.98 (0.97-0.99) | 1.00 (0.99-1.01) | 0.99 (0.97-1.00) | 1.16 (1.15-1.17) | 1.12 (1.11-1.13) |
| Race (Blacks vs. non Black) | 3.41 (3.15-3.70) | 3.43 (2.95-3.99) | 0.74 (0.66-0.83) | 0.56 (0.45-0.70) | 1.11 (1.01-1.21) | 0.80 (0.68-0.94) |
| HbA1c | 1.17 (1.10-1.23) | 1.23 (1.16-1.32) | 1.26 (1.18-1.34) | 1.26 (1.18-1.33) | 1.16 (1.10-1.22) | 1.19 (1.12-1.27) |

Race was significantly associated with hyperfiltration only for eGFR and eGFRr, but the association was borderline for the crude regression of absolute eGFRr (OR=1.11, 95% CI 1.01-1.21). According to standard eGFR, the risk of hyperfiltration was increased for Black participants (adjusted OR=3.43, 95% CI 2.95-3.99). However, when eGFR was calculated without the race-specific coefficient the risk of hyperfiltration was lower for Black compared to non-Black individuals in both crude and adjusted models: OR=0.74 (95% CI 0.66-0.83) and OR=0.56 (95% CI 0.45-0.70).

With the standard equations the risk of hyperfiltration for Black individuals was higher than in non-Black subjects across all BMI groups, although the magnitude of the association decreased at higher BMI (Supplementary table 1). When eGFR was calculated without the race-specific coefficient, Black race was significantly associated with reduced risk for hyperfiltration in the overweight and obese groups, (OR=0.7 95% CI 0.54-0.89, and OR=0.47 95%CI 0.37-0.60 respectively). The risk of hyperfiltration was identical between racial groups for the normal weight and overweight according to absolute eGFRr (OR=1.25 95% CI 0.99-1.57, OR=1.03, 95% CI 0.85-1.24 respectively), and the closest for the obese, when compared to the other two formulae (OR=0.8 95% CI 0.7-0.91).

Bland Altman analyses were performed to test the agreement between creatinine clearance, and each formula; eGFR, absolute eGFRr and absolute eGFRr across different BMI groups for Black individuals. There was a positive bias for all formulae across the complete cohort (supplementary figure 1).

Pearson’s coefficient was calculated to assess the correlation between each formula and creatinine clearance. Agreement between creatinine clearance and absolute eGFRr was better (r=0.64, p-value<.0001) than for clearance with either eGFR or eGFRr (r=0.52, p-value<.0001). Of note, removal of the race coefficient did not improve agreement between estimating formula and measured clearance.

## Discussion

We examined the incidence of hyperfiltration according to eGFR and eGFRr in a large cohort of adolescents to understand the effect of the race coefficient on prevalence of ORG.

In the complete cohort estimation with the standard formula translated to increased risk of hyperfiltration among Black individuals. However, after Removal of the race coefficient, rates of hyperfiltration among black adolescents were lower compared to non-Black adolescents in the overweight and obese groups and in adjusted analyses Black individuals were significantly less likely to have hyperfiltration. Furthermore, in individuals with 24-hour urine collections, estimates of creatinine clearance were identical in Black and non-Black individuals for every BMI group except the underweight, suggesting that the estimates including the race-specific coefficient were likely misclassifying hyperfiltration. The closest estimation of GFR to clearance was achieved by combining removal of the race coefficient with adjustment to individual BSA.

Obesity may increase the risk of CKD by extenuating other components of the metabolic syndrome (such as hypertension and insulin resistance) or by inducing glomerular hyperfiltration. Hyperfiltration rates ranged between 9-11% in a population of 2043 Black adults, mean age 52, over a follow up of 8 years. These rates are lower than in our cohort, probably reflecting longer duration of obesity and progression of hyperfiltration to overt focal segmental glomerulosclerosis (FSGS) in adults(24). Hyperfiltration rates in an NHANES cohort were increased in white adolescents (9.8%) compared to Black adolescents (7.4%), yet the authors did not report BMI in their population, questioning the relevance of this finding to obesity(25). In another cohort of 1575 teenagers BMI was positively correlated with hyperfiltration but information on race was not available(26). To our knowledge, our study is the first to jointly evaluate the relative contribution of race, and obesity, to the risk hyperfiltration in adolescents.

Given the discrepancy with measured creatinine clearance, it appears that the standard equation over diagnoses hyperfiltration in Black individuals as the incidence of hyperfiltration was higher in every BMI category. In contrast, removing the race coefficient from the estimation of GFR in young women, reversed the inter-race differences in hyperfiltration rates for obese and overweight individuals.

Presumably, the race coefficient should be removed from equations estimating GFR for the entire Black population since there is no biological basis for the social construct of “race”(27). Different cohorts and equations have suggested different race coefficients, depending on GFR range of participants and percent of Black subjects included(28)(29–31)(32). Of note, the same race corrections stemming from American Blacks failed to estimate GFR of other Black populations around the world(33). Yet, removal of the race coefficient alone, as suggested by some authors(34–36), did not achieve resemblance of eGFR between races and may also lead to underdiagnosis of ORG (figure 2), masking subsequent renal damage while reclassifying some teenagers as CKD.

A few previous studies investigated the association between baseline hyperfiltration status, and subsequent GFR decline. In a population-based outpatient data set(38) higher estimated GFR (eGFR) was associated with increased risk for doubling in serum creatinine level during a median follow-up of 35 months. Two other studies found that higher baseline eGFRs predicted a steeper eGFR decline in large community cohorts (39,40). A 19-year follow-up study in patients with type 1 diabetes found that higher baseline measured GFR was associated with more rapid decline in GFR (41).

Our analysis reveals a major caveat of the currently recommended formulae to estimate GFR. As modern medicine aspires precision and individualized approach, GFR estimation and CKD management lags behind, remaining standardized and general. The use of a unanimous BSA for the entire population obscures individual risk factors for renal morbidity. In order to reduce the burden of CKD for the black population, we should focus on evaluating changeable cardiometabolic risk factors such as obesity.

Our analysis has important limitations. GFR was not measured directly using inulin or radioisotope methods which are considered the gold standard measures of renal function(42,43). Clearly, the use of these exogenous markers to estimate GFR is impractical in clinical practice; and clinical trials evaluating measured GFR do not focus on participants with normal range kidney function. In addition, our data came from adolescents in a single urban area, and the results should be generalized cautiously to other populations.

Our investigation has some obvious strengths. We explored the interaction between race and obesity in a relatively healthy young population free from confounding effects of other co-morbidities potentially causing hyperfiltration such as hypertension and diabetes. Moreover, although our data came from a single urban area, the population was large and diverse and came from multiple health care institutions.

The higher than expected prevalence of renal morbidity among Black individuals may be attributed to the combination of lower socioeconomic status(44,45), mistrust toward medical establishment(46) deterring timely evaluation and treatment, and a flawed method of GFR estimation among other causes. In the very diverse and heterogenous group of Black individuals more attention should be given to the individual risk profile rather than to the common characteristics, that do not influence physiology such as race. Unique effects of traditional risk factors, such as obesity, that are reversed by awareness, and life style modification could profoundly influence disease progression and end organ damage.

In conclusion, poor healthcare outcomes aggregate unequally. As long as uncertainty persists about the effect of racial differences on renal morbidity, we should favor practices that may alleviate health inequities over those that may exacerbate them(47).

Our data suggests that one size fits all approaches to eGFR estimation should be avoided and that eGFR equations should be revised to reflect differential risk and avoid over standardization that groups heterogenous populations under similar categories.

## Data Availability

Data have been provided by the INSIGHT Clinical Research Network and can be made available upon request. The
INSIGHT Governance Board will review each request.

## Abbreviations

BSA: body surface area
ORG: Obesity Related Glomerulopathy
GFR: Glomerular filtration rate
CrCl: Creatinine clearance

