## supplementary material for "Removal of the race coefficient and adjustment to individual BSA provide the most accurate estimation for GFR in Black adolescents"

**Dana Bielopolski^1^, Ohad S. Bentur^1^, Neha Singh^1^, Roger D. Vaughan^1^, David M. Charytan ^3^, Rhonda G. Kost^1^, Jonathan N. Tobin^1,2^**

**^1^**The Rockefeller University Center for Clinical and Translational Science, New York NY

^2^Clinical Directors Network (CDN), New York NY

^3^Nephrology Division, New York University Langone Medical Center and Grossman School of Medicine, New York, New York

Supplementary table 1 - **Odds ratio for hyperfiltration of Black individuals vs. non-Black individuals across BMI groups**. Hyperfiltration was calculated according to eGFR (KDIGO reccomended formula), eGFRr (KDIGO reccomended formula for age without the race coefficient) and absolute eGFRr (KDIGO reccomended formula for age without the race coefficient and adjusted to individual BSA).

|  | underweight | Normal weight | Overweight | obese |
| --- | --- | --- | --- | --- |
| eGFR | 3.09 (2.20- 4.35) | 3.92 (3.50- 4.39) | 3.57 (3.00- 4.25) | 2.50 (2.12- 2.95) |
| eGFRr | 1.16 (0.76-1.74) | 0.87 (0.75-1.02) | 0.70 (0.54-0.89) | 0.47 (0.37-0.60) |
| Absolute eGFRr | 4.26 (1.138-13.14) | 1.25 (0.99-1.57) | 1.03 (0.85-1.24) | 0.80 (0.70-0.91) |

Supplementary figure 1 –**Bland Altman plot for the different formulae vs. clearance in Black individuals** (N=102). Panel A - eGFR vs. clearance, panel B - absolute eGFRr vs. clearance, panel C – eGFRr vs. clearance.

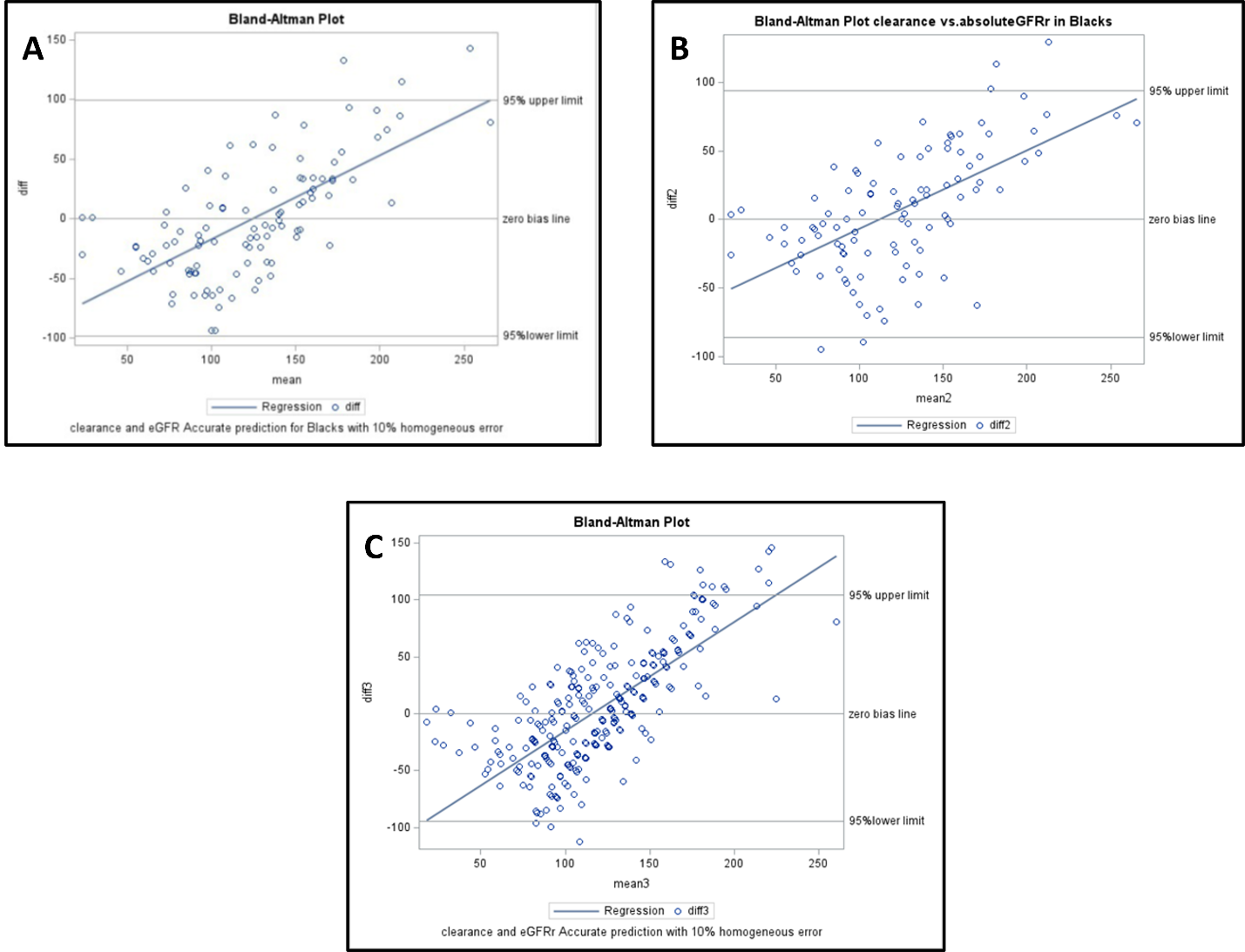
